# Early pancreatic tissue necrosis diagnoses by perfusion CT; the study K-Pa

**DOI:** 10.1101/2024.01.13.24301278

**Authors:** Guillaume Herpe, Claire Renaud, Jean-Pierre Tasu

## Abstract

**PURPOSE:** Acute pancreatitis (AP) is associated with high mortality and morbidity rates in case of necrotic forms. Risk assessment should be early performed to stratify patients into higher- and lower-risk of severe form to assist triage. In severe pancreatitis, capillary permeability increases, thereby contributing to capillary leakage which explains organ failures and or tissue necrosis. The aim of this study was therefore to evaluate pancreatic permeability by perfusion CT (pCT).

**METHODS:** From March 2018 to November 2018, patients with suspected AP and who underwent CT at admission were prospectively included. AP cases were classified according to the revised Atlanta classification. A permeability parameter, called k-trans, was measured from pCT in 3 pancreatic areas (normal parenchyma zone, defined by an area of normal CT pattern, pathological zone, defined as an area of parenchymal enlargement and or lack of enhancement and an intermediary zone defined by an area between normal and pathological areas) by to two observers. K-trans values in necrotic and interstitial forms for each zone were compared. To estimate reproducibility of the measure, inter-observer and intra-observer agreement was evaluated by a Bland and Altman test.

**RESULTS:** 15 patients were enrolled (mean age 45.50 years old, +/-17.70). Four acute pancreatitis were necrotic, and 11 interstitial.

Mean k-trans in pathologic zone of necrotic forms was significantly lower (mean=0.08) than in interstitial (mean=0.53), p= 0.0003. In both forms, k-trans values were significantly lower in pathologic zones than in intermediary and normal zones and the higher k-trans values were obtained in intermediary zones. Intra-observer reproducibility was good. Inter-observer reproducibility was acceptable, one bias was reported, close to zero (−0.052) with limited statistic difference.

**CONCLUSION:** K-trans parameter, a well-known marker of tissue permeability, can be estimated by pCT. This parameter seems to be linked to local necrosis and could be used as a discriminant mean to diagnose necrotic from interstitial types of AP in the early phase of disease.

## INTRODUCTION

Acute pancreatitis (AP) is one of the most common diseases of the gastrointestinal tract, with an incidence varying between 4.9 and 73.4 cases per 100,000 worldwide (1). Two distinct phases of AP have been described by the revised Atlanta classification (2): an early one, within 1 week, characterized by the systemic inflammatory response syndrome (SIRS) and/or organ failure (defined by a modified Marshall Score) and a late one (after 1 week), characterized by local complications. In addition to organ failure, pancreatic necrosis, including parenchyma and/or extra pancreatic involvement, is one of the main criteria associated with complications with a death rate of around 30% (3). A large majority of patients presents a mild AP form associated with uncomplicated course. However, in patient failing to improve after 48–72 hours, computed tomography (CT), or MRI imaging is recommended to assess local complications such as pancreatic necrosis (4). However, there are two issues on necrosis diagnosis; 1-The CT delay postpones the risk assessment of local complication although this risk should have been early performed to stratify patients to assist triage at admission; 2-For detecting pancreatic parenchymal necrosis, interobserver agreement is moderate (κLJ=LJ0.45) (5) without any improvement with time.

Perfusion CT (pCT) have already been exhaustively studied in AP (3, 6–11). Assessment of pancreatic necrosis development is possible at the early stage of AP (7, 12) using pancreatic blood flow (BF) or median blood volume (BV). However, there are a main limitation in using these parameters; BF and BV corresponds to the volume of capillary blood contained in a certain volume of tissue. These parameters reflect mostly the early intrapancreatic protease activation and acinar cell injury causing a local and systemic inflammatory (13). In another ways, it has been shown that endothelin plays a major role in mediating pancreas capillary permeability. In severe pancreatitis, capillary permeability increases even outside the pancreas, thereby contributing to capillary leakage and could explain organ failures and or necrosis. (14). However, to date and to our knowledge, this parameter has never been measured in human AP. Therefore, the aim of this pilot study was to assess the ability of pCT) to measure pancreatic capillary permeability, estimed by the k-trans, in cases of AP.

## METHODS

### Patients

From March 2018 to November 2018, a prospective study from Poitiers Universitary Hospital was conducted including patients over 18 y.o. underwent CT for acute pancreatitis suspicion.

Diagnosis of AP was established according to 2012 revised Atlanta Classification criteria REF including clinical and biological parameters. Two of the following 3 features were required: 1) abdominal pain consistent with acute pancreatitis (epigastric pain radiating through the back), 2) a 3 times increased amylase or lipase, 3) signs of acute pancreatitis on the contrast-enhanced computed tomography.

The onset of AP, according to the revised Atlanta Classification, was defined at the time of onset of abdominal pain.

The exclusion criteria of this study were age under 18 years old, pregnancy and iodinated contrast medium injection contraindication (history of reaction to iodinated contrast agent or severe renal failure).

The patients signed a consent paper after receiving an oral and written information about the study. This study was approved by Ethic Comity (ID-RCB 2017-A02708-45).

The following data were recorded; age, gender, reason of admission at the Emergency Department (ED), date of the onset of symptoms, medical or surgical history, intensity of pain using standardized visual analog scale (ranging from 0 to 10), size, weight, serum lipase at admission, etiology of AP, complications.

### pCT protocol

pCT examinations were performed on one unique device (TOSHIBA Aquilion 64, Tokyo, Japan) according to a standardized protocol including; 1-A non-contrast phase, (2 mm slice thickness, 512x 512 matrix, 100kV, modulation mAs in the z direction, pitch 1.4), 2-a perfusion phase, performed focused on the pancreas after injection of 40 mL of iodinated contrast agent (IOMERON® 400mgI/mL at a rate of 6 mL/second using an automatic contrast injector (MEDTRON ACCUTRON CT-D (n°861013837)) via a cubital vein. The dynamic acquisition started simultaneously with the injection and consisted in 64 slices of 0.5 mm every 3 seconds during 90 seconds. The parameters were; matrix 512 x 512, thickness 2 mm, 100 Kv and 80 mAS, pitch1,4); 3-Two more phases were performed, one 35 seconds (pancreatic phase) and 80 seconds (portal phase) after a second intra venous injection of 60 mL of iodinated contrast agent (IOMERON® 400mgI/mL at a rate of 3 mL/second using the same automatic contrast injector (MEDTRON ACCUTRON CT-D (n°861013837). Patients were required to breathe quietly during examinations.

### Image analysis

After the pCT images were acquired, different radiologists, with 3 or more years of experience made prospective diagnoses of acute pancreatitis on PACS station (Maincare V12, Vancouver, Canada). This diagnosis was performed on the date of admission in the Emergency Department. The radiologists had full access to clinical information. The morphological analysis was based on revised Atlanta Classification identifying three forms of acute pancreatitis:

– the interstitial edematous form, characterized by a pancreatic diffuse or local enlargement with normal enhancement and those with no morphological change.

– the necrotic ones, based on a glandular lack of enhancement of enhancement of the gland of 30 Hounsfield Unities (H.U.) in the portal phase. This definition was used to have a standardized rule for necrosis diagnosis.

For simplicity of presentation, interstitial edematous forms and morphologically normal forms were grouped and called the non-necrotic forms (written 0 in the results), in opposite to the necrotic ones (written 1 in the results).

For the study, pCT images were re-analyzed on a dedicated software (OLEA SPHERE ®, Paris, France). Permeability parameters were calculated using Extended Tofts model; the Extended Tofts model is a two-compartment model based on agent contrast diffusing from intra-vascular space (IVS) to the extra-vascular extra-cellular space (EES). The permeability parameter studied in our study was k-trans, a permeability surface area product per volume, from IVS to EES, expressed in min^-1^.

To estimate pancreatic k-trans, two investigators drawn manually six regions of interest (ROI), avoiding vessels and ducts, with an average surface of 25 mm^2^ placed on the pancreas gland. The pancreas was divided in three different zones in which 2 measures were perfomed:

– The normal zone (N), where the pancreatic gland strictly showed no morphological change.

– The pathological zone (P), where the pancreatic showed any modification such as lack of enhancement in the necrotic form or parenchymal enlargement.

– The intermediary zone (I), also morphologically normal, located at least 10 mm border zone of the pathological zone.

Numerical values of k-trans, rBV (relative blood volume) and color-coded parametric maps of k-trans were obtained (figure 1). Mean value was computed for each zone.

**Figure 1:**
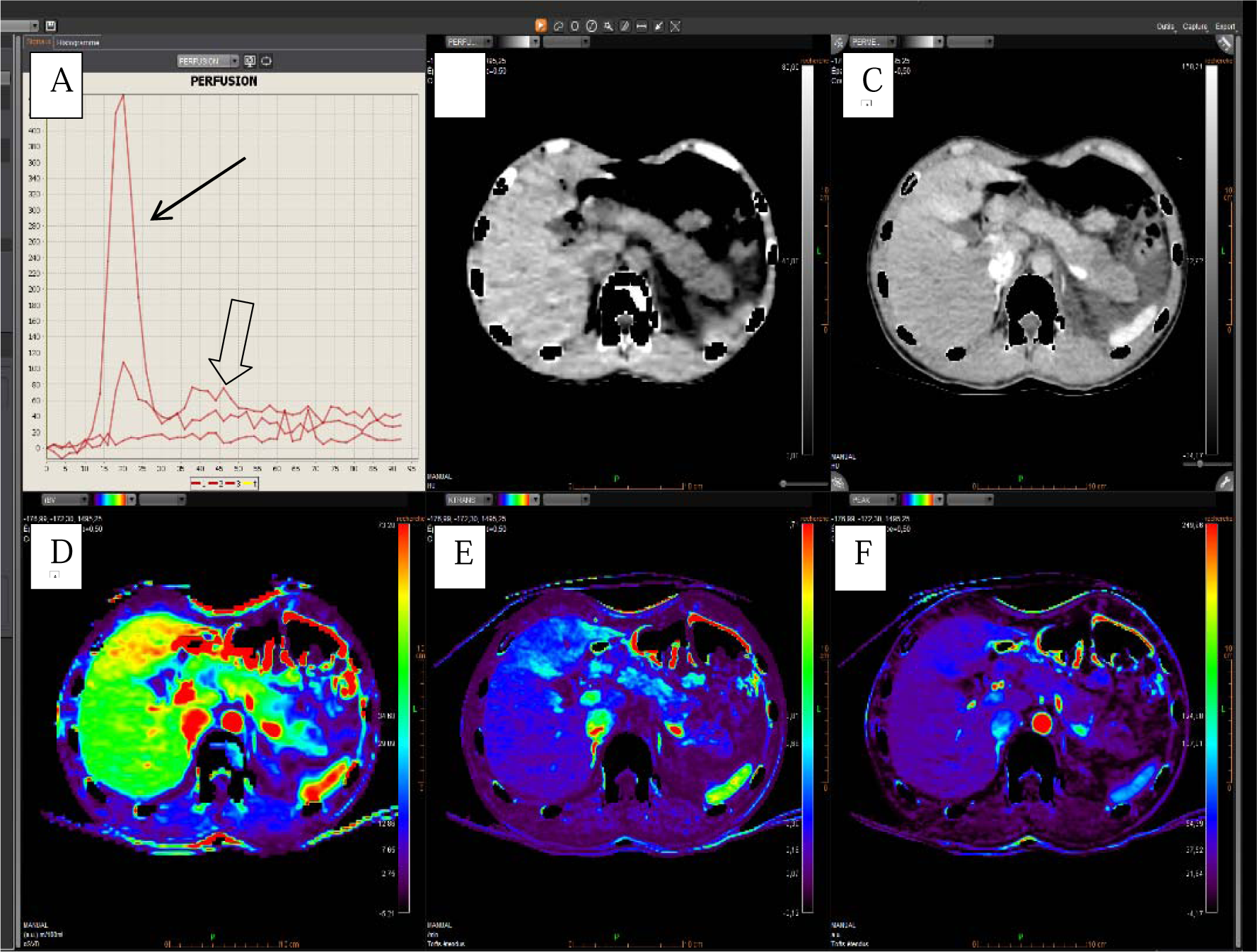
Perfusion curve, pCT images and colour perfusion maps for k-trans and rBV. A: perfusion curves estimate in aorta and pancreatic tissue (respectively black arrow and open arrow); B and C: native CT perfusion before and after injection of iodine contrast; D:??? E: F; ???

The first investigator repeated the analysis twice and the second investigator once to assess intraobserver and interobserver reproductibilities by Bland-Altman method.

A total of 270 k-trans values were obtained with this protocol by the two investigators, repeating 2 ROI for each defined zone: 72 in the necrotic group, 198 in the non-necrotic group. Figure 2 illustrates image of k-trans measurement.

**Figure 2.**
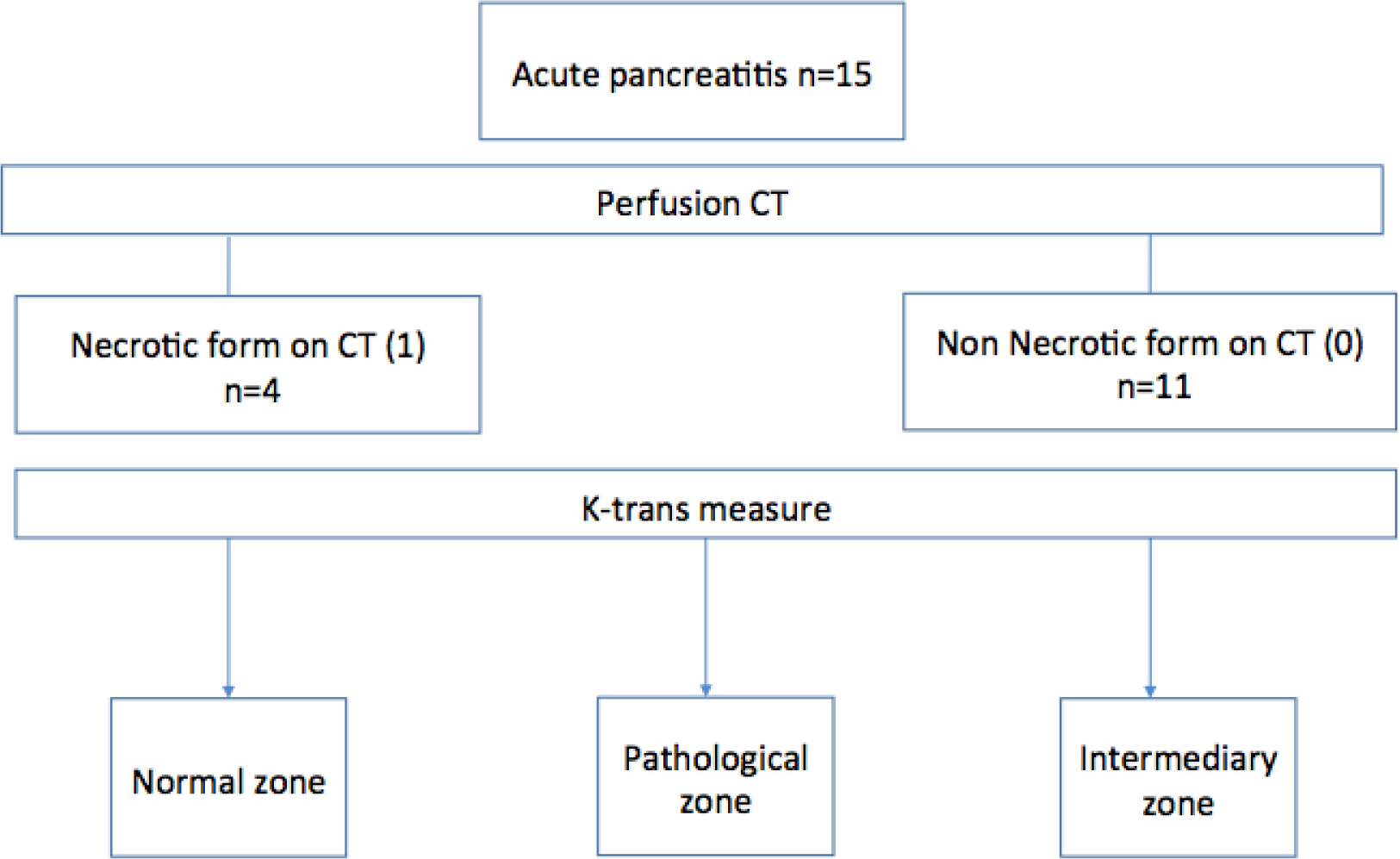
Flow chart

#### Radiation dose

Radiation dose exposure was computed using a dedicated dose management software (Radimetrics^TM^ Entreprise platform, Bayer Healthcare, Whippany, NJ, USA).

#### Statistical analysis

Descriptive data are presented as mean ± standard deviation for quantitative data, numbers and proportions for qualitative data. The variable distribution was checked for approximately normal distribution. Differences between groups were estimated and tested using repeated measures ANOVA and least-squares adjusment. Inter-observer agreement among two radiologists and intra-observer agreement were evaluated per zone using the Bland-Altman method. Biais significance was evaluated using the Student t test. P-values of less than 0.05 were deemed significant. SAS 9.4 software (SAS Institute Inc, CARY NV, USA) was used for the statistical analysis.

## RESULTS

### Patients

We enrolled 15 patients with AP (11 males and 4 females with an average age of 45.5, 21-89 year-old); figure 2 gives the flow-chart of the study. The clinical and biological characteristics are presented in Table 1. All patients were admitted for abdominal pain. Mean delay between admission and pCT was 06 hours 20. All patients with necrosis forms had their CT less than 24 hours from the onset of symptoms (from 5 hours to 14 hours).

**Table 1.**
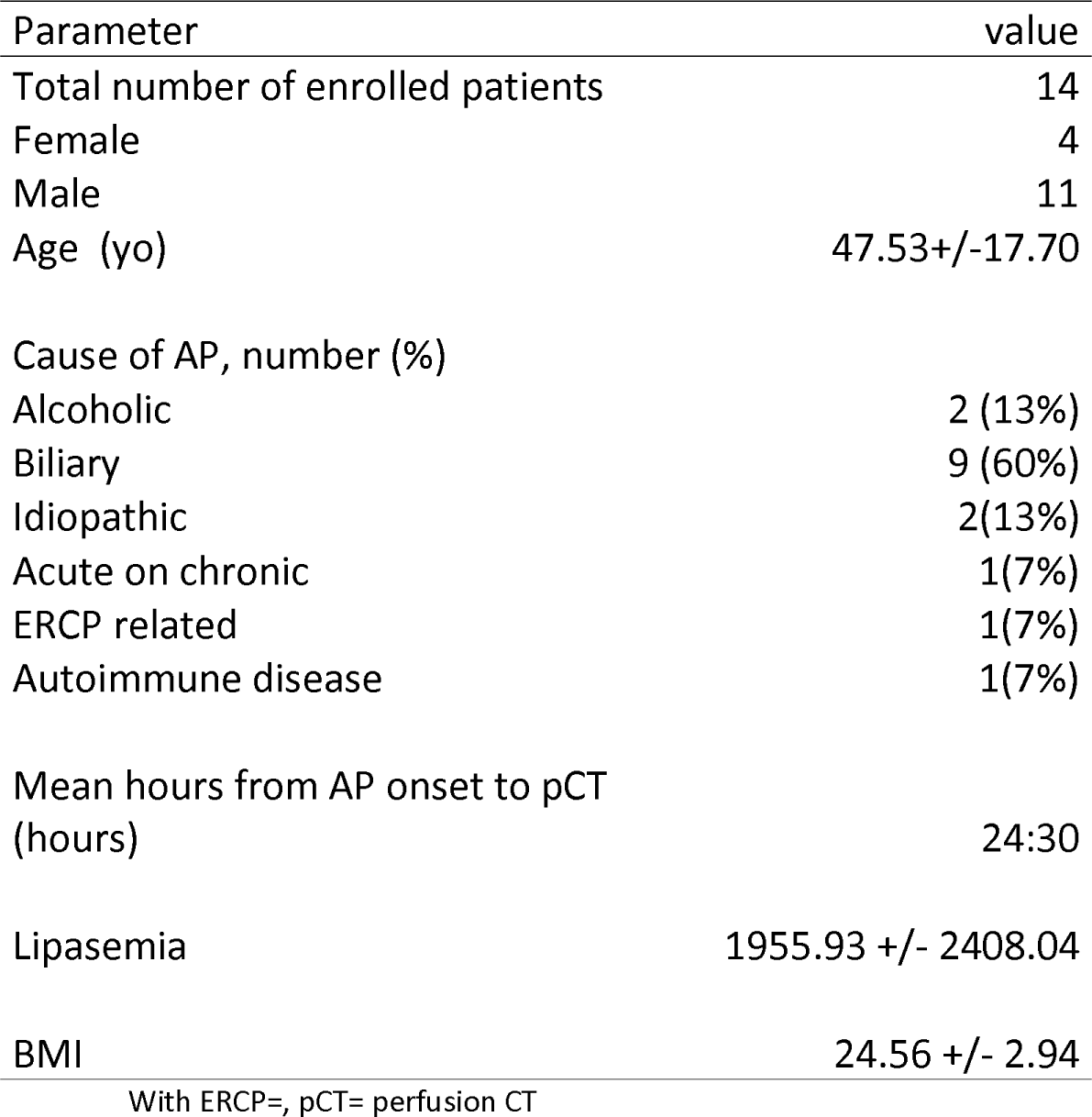
Characteristics of the enrolled patients.

Causes of AP were cholelithiasis in 9 patients (60%), alcohol in 2 patients (13%) and autoimmune disease for 1 patient (7%). One patient developed the disease after a retrograde cholangiopancreatography. Two forms were diagnosed as “idiopathic” considering the negative research for cause; one autoimmune cause was suspected but not eventually proved. Mean duration of Hospital stay was 3.64 days± 2.06 with 10 patients admitted in Gastroenterology and Hepatology Department, 2 in post-emergency Unit, 3 in Intensive care. Two pulmonary infections were observed. One patient died due to multi-organ failure.

According to Atlanta classification, 5 presented a mild AP, 6 a moderate one and 4 a severe form. SIRS was present for 7 patients on the first day of admission. Patients were classified in two groups: necrotic form, called “1” in the results (n=4, 26.5%) interstitial edematous pancreatitis form, called “0”, including those with parenchyma edema (n=7, 47%) and those with no morphological modification (n=4, 26.5%).

#### Perfusion and permeability analysis

K-trans values are given Table 2 and the most important results are reported Table 3. Mean values of rBV are shown Table 4.

**Table 2.** Mean k-trans values according to forms and zones.

| Forme | Zone | n | Moyenne | SD | Mini | Max |
| --- | --- | --- | --- | --- | --- | --- |
| Interstitial | I | 66 | 0.61 | 0.23 | 0.2 | 1.04 |
|  | N | 66 | 0.6 | 0.21 | 0.2 | 1 |
|  | P | 66 | 0.53 | 0.21 | 0.12 | 0.91 |
| Necrotic | I | 24 | 0.59 | 0.21 | 0.29 | 1 |
|  | N | 24 | 0.42 | 0.2 | 0.18 | 0.75 |
|  | P | 24 | 0.08 | 0.03 | 0.02 | 0.14 |

*With; I: Intermediary; N: Normal; P: Pathological; n: number of values obtained; SD: Standard Deviation; mini: minimum; max :maximum*

**Table 3.** Comparison of mean k-trans values according to forms and zones: Last mean squares results.

| form/zone | mean ktrans | form/zone | mean ktrans | Estimation | p |
| --- | --- | --- | --- | --- | --- |
| N/Pathological | 0.08 | N/Normal | 0.42 | 0.3363 | p<0.0001 |
|  | 0.08 | N/Intermediary | 0.59 | 0.5129 | p<0.0001 |
| N/Pathological | 0.08 | I/Pathological | 0.53 | 0.4456 | p=0.003 |
| I/Pathological | 0.53 | I/Pathological | 0.6 | 0.0759 | p=0.0011 |
|  |  | I/Intermediary | 0.61 | 0.086 | p=0.003 |

With; I: interstitial form; N: necrotic form;
Pourquoi 2 lignes N???

**Table 4.**
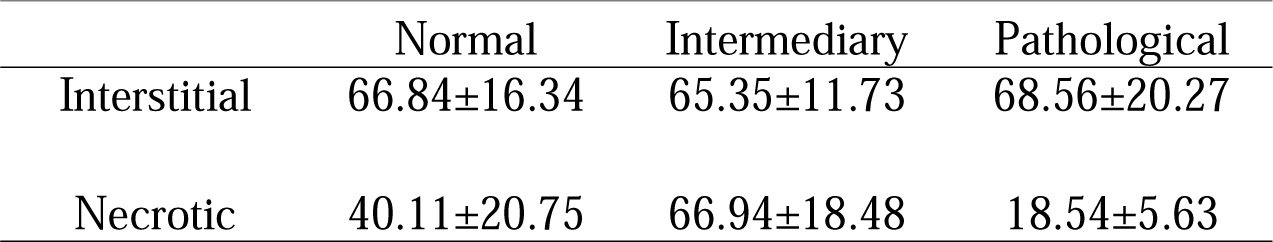
Means values of rBV according to zones and forms.

|  | Normal | Intermediary | Pathological |
| --- | --- | --- | --- |
| Interstitial | 66.84±16.34 | 65.35±11.73 | 68.56±20.27 |
| Necrotic | 40.11±20.75 | 66.94±18.48 | 18.54±5.63 |

K-trans in pathologic zone of necrotic forms was significantly lower (mean=0.08±0.03) than in non necrotic forms (mean=0.53±0.21), p= 0.0003. In both forms, k-trans values were lower in pathologic zones than in Intermediary and Normal zones: Among necrotic forms, k-trans in pathologic zone (mean=0.08±0.03) was significantly lower than in Normal zone (mean=0.42±0.2), and Intermediary (mean=0.59±0.21), p<0.0001. Differences were lower in the non-necrotic forms but remained significant: p= 0.0011 in zone N (mean k-trans in normal zone=0.60±0.21) and p= 0.0003 in Intermediary zone (mean k trans = 0.61±0.23)

Figure 3 illustrates the differences between mean k trans according to the zone in non-necrotic and necrotic form.

**Figure 3.**
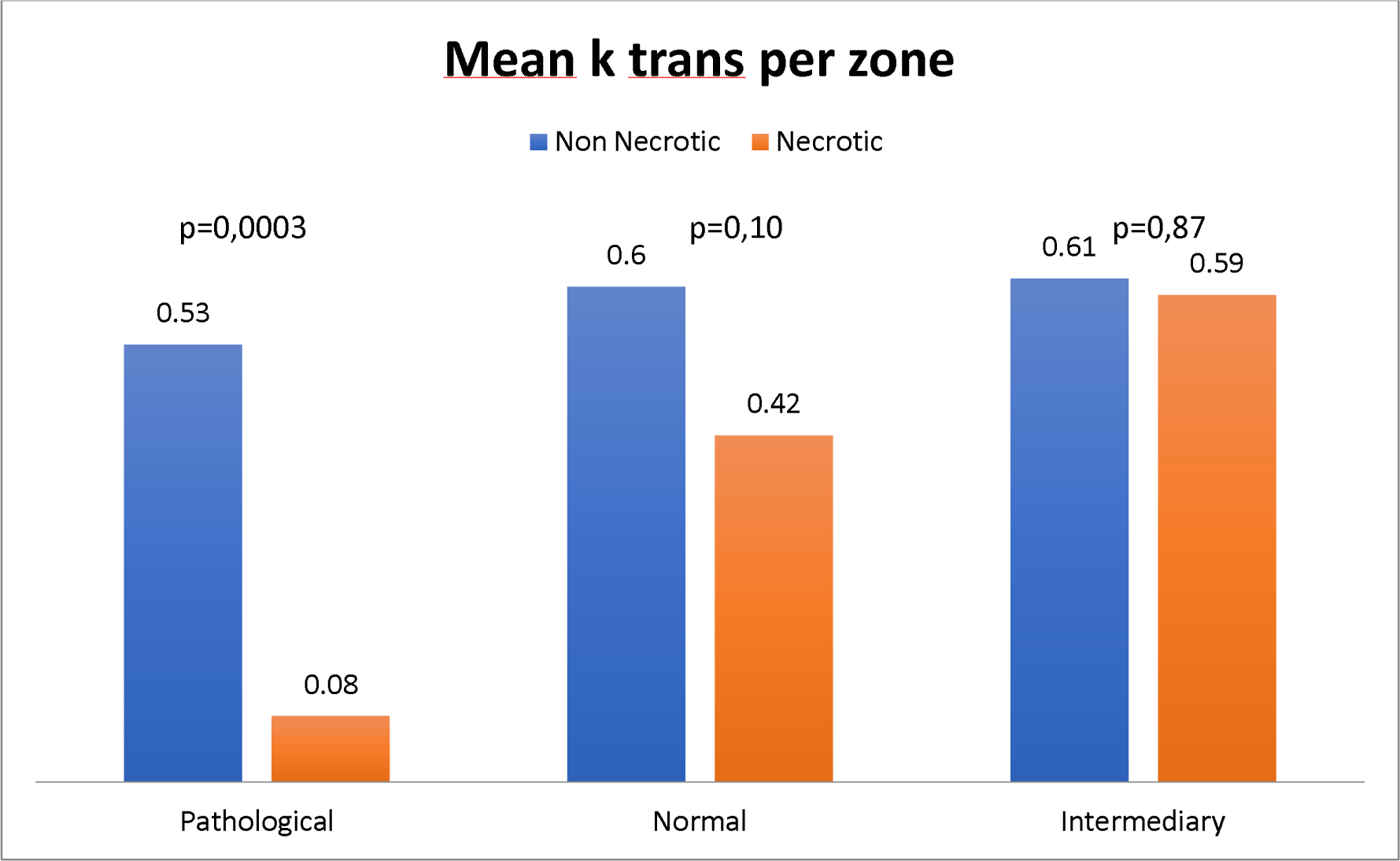
Differences between mean k-trans according to the glandular zone in necrotic and non necrotic forms.

The higher k-trans values were obtained in Intermediary zones for both acute pancreatitis form: mean k-trans in Intermediary zones in Necrotic forms was 0.59±0.21 and 0.61±0.23 in non necrotic forms, p=0.8655.

For each zone, regardless of the acute pancreatitis form, inter-observer and intra-observer agreement was studied by the Bland-Altman method and reported in Figure 4. The first observer values are written A and B as he repeated the measurement twice. The second observer results are written C and were compared to results A. Intra-observer evaluation did show good repeatability. In the pathological zone, the 95 per cent limits of agreement ranged from −0.170 to 0.173. In normal zone, the 95 per cent limits of agreement ranged from −0.139 to 0.146. One bias was reported in inter-observer evaluation, very close to zero (−0.052) with limited statistic difference, p= 0.05, in normal zone. In the pathological zone, the 95 per cent limits of agreement ranged from −0.290 to 0.307.

**Figure 4.**
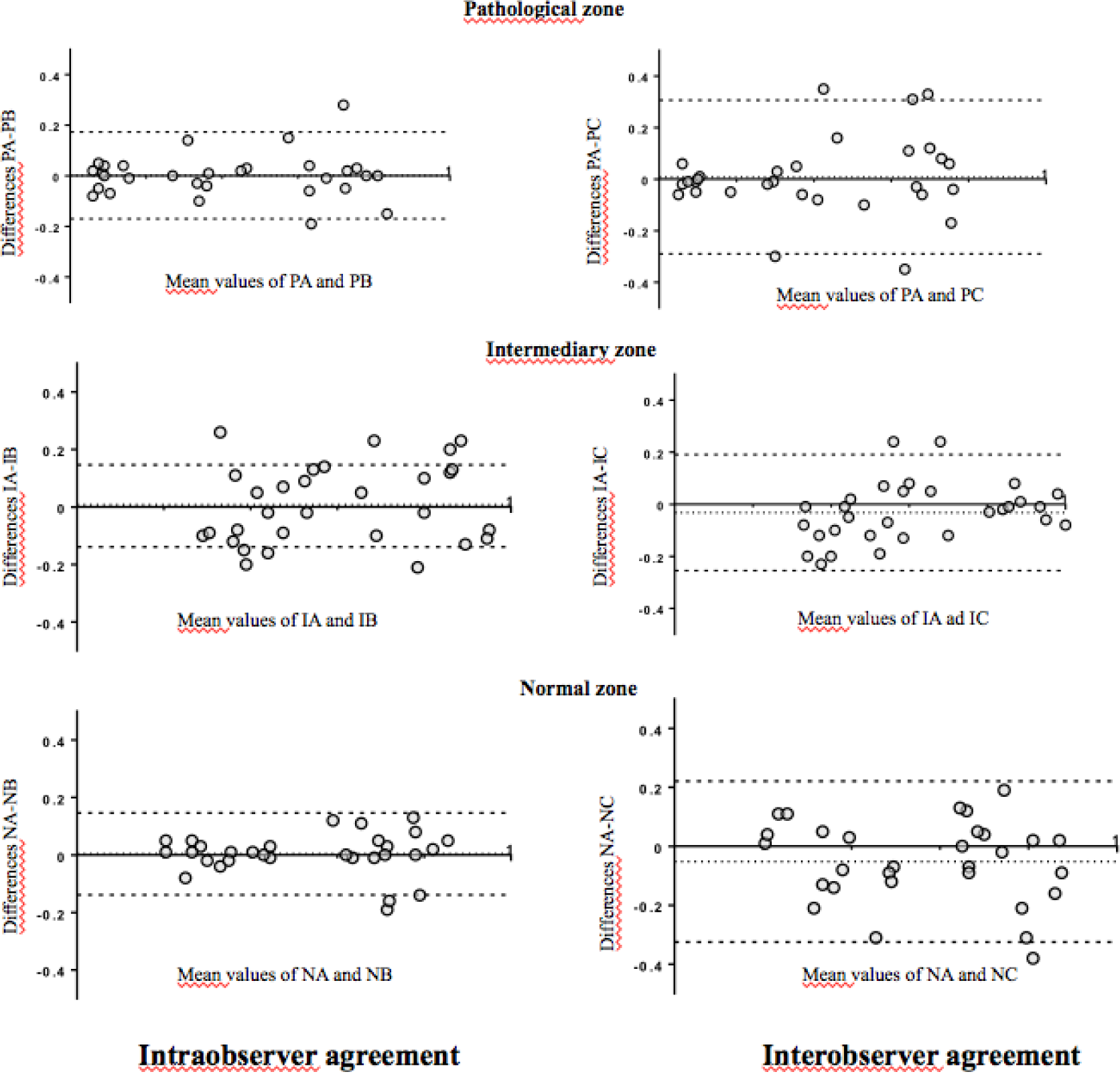
Bland and Altman plot: Intraobserver and interobserver agreement. *With; A and B: first observer; C: second observer, P: Pathological, I: Intermediary, N: Normal*

#### DOSE RADIATION

The mean dose of dynamic acquisition was 562 mGy.cm^-1^ (range 437 to 582 mGy.cm^-1^). Mean total dose per patient was 1628.86 mGy.cm^-1^.

## DISCUSSION

This preliminary study shows that pCT can estimate a capillary permeability parameter without any overcost in terms of radiation or iodine load. A total of 15 patients was enrolled. The permeability parameter assessed in this study, the k-trans, showed lower values in pathologic zones, with significant difference according to the form and higher values in intermediary zones.

They are 2 main points related to AP severity; 1-the systemic inflammatory response syndrome (SIRS) which predicts severe acute pancreatitis at admission and during the first 48 hours; 2-the necrosis form of AP. Increase of capillary permeability could explain both (13). By the way, evaluation of capillary permeability is a great challenge and this preliminary study is the first one showing that pCT could be used for.

Pancreatic but also extra pancreatic capillary permeability increasing is likely one of the major parameters explaining severity of AP; fluid loss into the third space and reduce of capillary blood flow in the pancreas could explain organ failure and local parenchyma necrosis. Some authors even suggest that severe AP is more than a local disease, a systemic microcirculatory dysfunction syndrome (15, 16). The higher k-trans mean observed into the intermediary zone could reflects local higher capillary permeability leading to fluid loss into the third space and systemic inflammatory response. A previous study has shown that perfusion can predict of organ failure; the area under the curve (AUC) for predicting organ failure was between 68 and 74 according to the readers (3). However, blood flow and blood volume are not related to permeability. The authors explained therefore that these parameters, increased in cases of AP, could reflect mostly local inflammation with early intrapancreatic protease activation and acinar cell injury, which cause a local and systemic inflammatory response due to proinflammatory cytokines and secondary mediators released (13). However, direct measurement of local capillary permeability should be a key-point to better understand this complex pathology. Our preliminary results are encouraging but should be confirmed by further studies.

Pancreatic severity is also associated with necrosis and it has been already shown that perfusion imaging can predict necrosis (7–10). Pancreatic necrosis can be underestimated within 2-3 days after the onset of symptoms but is a sign of evolving pancreatic disease taking few days to manifest on imaging as it’s illustrated on figure 5. Hence conventional CT too early perform could neither diagnose pancreatic necrosis accurately nor estimate its extent (17) and CT necrosis detection accuracy increases with after 3 days (2); this delay leads to postpone suitable aggressive treatment and intensive care which can affect the overall prognosis of some patients. Some previous reports have already demonstrated interest of pCT in this condition (7–10). Based on receiver operating curve (ROC) analysis, the sensitivity and the specificity of pCT could even reach 100% and specificity 100% for predicting necrosis (7). Statistical differences in perfusion parameters were found between mild and severe forms.

**Figure 5.**
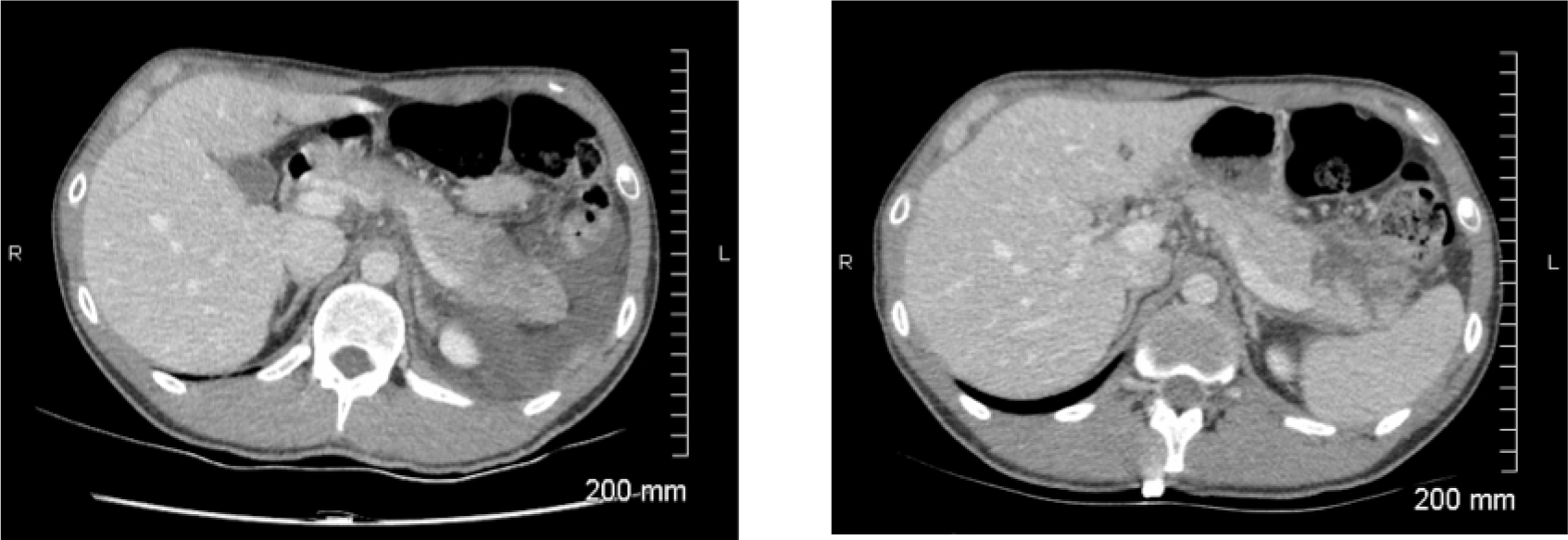
Comparison of lack of enhancement on pCT at admission and CT control processed 72 hours after admission in necrotic form.

(12). Our mean values of rBV are higher than those reported in these previous studies, which illustrate the variability for perfusion parameter measurements. The lower k-trans observed in the necrotic area is likely related to reduction of capillary flow. Regarding to the limited number of patients, we were not able to determine k-trans cut-off. Nevertheless, the significant difference between k-trans in necrosis and non-necrosis AP lead us to think that glandular zone with k-trans value < 0.1 min^-1^ should correspond to necrotic tissue. Nevertheless, this study is too preliminary and further studies are required to confirm interest of permability parameter to depict necrosis in addition of blood flow and blood volume.

Moderate interobserver agreement to categorize pancreatic and peripancreatic collections by CT in the first 4 weeks after symptom onset demontrastes that necrosis diagnosis can be challenging (5). Spectral CT, substraction CT or perfusion CT may improve agreement but there is a lack on data in the literature on reproducibility of these techniques. In our study, intra and inter-observer agreement for k-trans measurement seem to be reasonable: bias were very close to zero, only one was slightly significant (p= 0.05, in normal zone). In intra-observer evaluation did show good repeatability, all bias were very close to zero and no significant. For the intra-observer, in the pathological zone, the 95 per cent limits of agreement ranged from −0.170 to 0.173 which was entirely acceptable. The bias was not significant (0.001, p=0.93). We can therefore consider the k-trans measure as reproductible which is a real issue to diagnose necrosis by CT.

There are several limitations of this study; 1-it’s a pilot study and the number of patient is by the way limited; 2-Perfusion parameters were not correlated with angiographic findings; 3-We do not have normal k-trans value and establishing k-trans levels could be an interesting purpose of subsequent study; 4-No diagnosis reference for necrosis was obtained; routine aspiration would have been unethical and Magnetic Resonance Imaging (MRI) was not available in emergency in our center. Follow-up is not a perfect reference standard in this case. For instance, complete resolution of a collection after 4 weeks does not prove that the collection was not necrotic. 4-Perfusion parameters may vary according to CT device, mathematical model and software used and hence could not be comparable across centers without standardization.

## CONCLUSION

Perfusion CT is a tool to measure capillary permeability which could have an immense clinical importance to better understand AP diseases. This could lead to a better and early triage into a more aggressive line of management.

## Data Availability

All data produced in the present work are contained in the manuscript

## LIST OF ABBREVIATIONS

AP: Acute Pancreatitis
AUC: Area under the curve
BMI: Body mass index
CT: Computed Tomography
pCT: Perfusion Computed Tomography
rBV: Relative blood volume

